# The effects of sustained fitness improvement on the gut microbiome: A longitudinal, repeated measures case-study approach

**DOI:** 10.1101/2020.06.04.20046292

**Authors:** Wiley Barton, Owen Cronin, Isabel Garcia-Perez, Ronan Whiston, Elaine Holmes, Trevor Woods, Catherine B. Molloy, Michael G. Molloy, Fergus Shanahan, Paul D. Cotter, Orla O’Sullivan

## Abstract

**Objective:** The composition and metabolic function of the gut microbiome in the elite athlete differs from that of non-athletes. However, short-term fitness improvement in the sedentary adult does not replicate the microbiome characteristics seen in the athlete. Whether sustained fitness improvement over a prolonged period can lead to pronounced and beneficial alteration in the gut microbiome is unknown. The objective was to explore this possibility.

**Methods:** This study used a repeated-measures, case-study approach to explore changes in the gut microbiome of two unfit volunteers undertaking progressive exercise training over a 6-month period. Training was to culminate in the completion of a marathon or Olympic-distance triathlon. The volunteers were sampled every two weeks for six months and microbiome, metabolome, diet, body composition, and cardiorespiratory fitness data were recorded.

**Results:** Both participants completed their respective goals with improved body composition and fitness parameters over the training period. Increases in α-diversity of the gut microbiota occurred with sustained training and fluctuations occurred in response to training events (e.g., injury, illness and training peaks). Participants’ fat mass and BMI reduced during the study and was significantly associated with increased urinary measurements of N-methyl nicotinate (P value < 0.001) and hippurate (P value < 0.05), and decreased phenylacetylglutamine (P value < 0.05).

**Conclusion:** These results suggest that sustained fitness improvements result in alterations to gut microbiota and physiologically-relevant metabolites. This study provides longitudinal analysis of the response of the gut microbiome to real-world events during progressive fitness training, including intercurrent illness and injury.

## 1 Introduction

The available evidence suggests physical activity exerts a modest influence on the gut microbiome of individuals unaccustomed to exercise. This pertains to both the composition and metabolic production of the microbiota [1–5] (as reviewed [6–10]). Previous studies have highlighted the effects of exercise on the gut microbiota in mice [11] and lean and obese humans [4], over short-term exercise interventions (6 to 8 weeks). In contrast, professional athletes harbour a gut microbiome that is taxonomically and functionally distinct and more diverse than less physically fit individuals [12–18]. It is possible, as is believed in the elite athlete, that sustained improvements in cardiorespiratory fitness and body composition through exercise may lead to beneficial alterations in the characteristics of the gut microbiome that are associated with health and fitness (e.g., increased compositional alpha-diversity). However, the impact of longer-term exercise on the gut microbiota remains unknown [1]. Furthermore alterations in diet, that often occur in unison with exercise (be they intentional or unintentional), represent a possible confounding influence in the interpretation of studies examining the relationship between exercise and the gut microbiota.

To address this and to understand the relationship between sustained fitness improvement and the gut microbiome, we used a repeated-measures, longitudinal case-study method to detail changes in the gut microbiome over 6 months in two participants embarking on different training programs with specific training goals. Such study designs have been successfully implemented to interrogate longitudinal effects on the microbiome over time [19] and in fact have recently demonstrated that intense and prolonged ultra-endurance exercise leads to increased alpha-diversity in the gut microbiota [16]. This permitted a detailed interrogation of the subjects’ microbiome on a frequent basis over the training epoch, also facilitating the collection of detailed dietary information, allowing us to interpret exercise-related microbial changes with greater confidence. To add to our previous work in physically inactive individuals, participants with relatively low fitness levels at baseline were recruited [1]. To examine possible differences induced by training type, one volunteer combined regular resistance and aerobic training over 6 months, while the other focussed on aerobic conditioning alone.

## 2 Methods

### 2.1 Experimental model and ethical approval

An overview of the study design is shown in Figure 1. The study was approved by the Cork Clinical Research Ethics Committee (CREC), and was conducted in accordance with the Declaration of Helsinki. Both volunteers provided written informed consent before beginning study participation.

**Figure 1.**
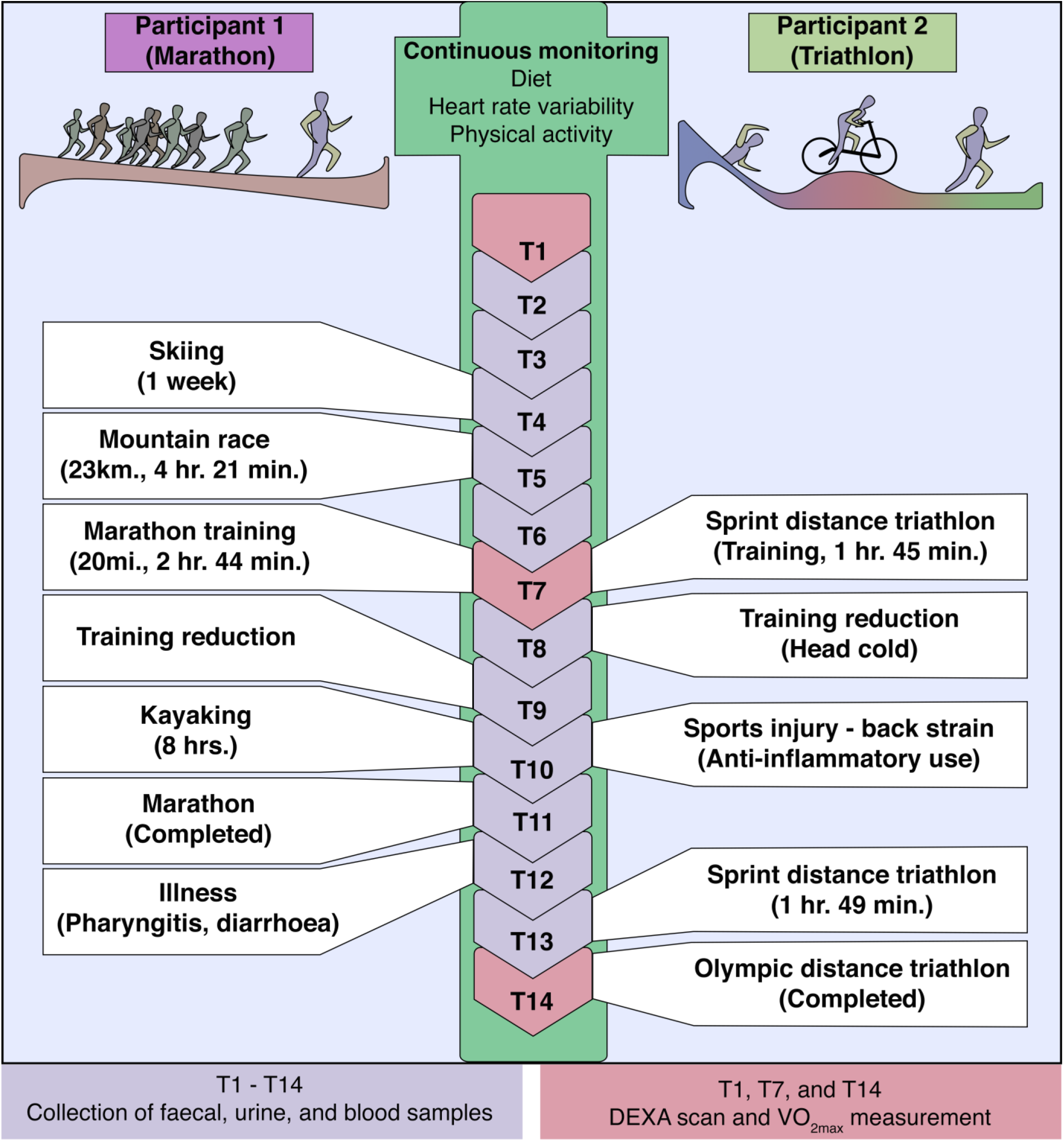
Study overview. The two participants supplied blood, fecal, and urine samples at every 2 week long time point. DEXA scan and measurement of VO_2max_ occurred at the beginning (T0) mid-point (T7) and final time point (T14). Diet, heart rate variability, and physical activity were continuously monitored. VO_2max_, estimated maximal oxygen consumption.

### 2.2 Study recruitment and safe participation

Two male participants were recruited from University College Cork. Both fulfilled inclusion and exclusion criteria (Table S1). Baseline levels of physical activity were assessed using the International Physical Activity Questionnaire (IPAQ) short form [20]. Safe participation in the study was ensured by medical screening of both participants using an adapted version of the safe participation questionnaire of the American College of Sports Medicine [21].

### 2.3 Exercise intervention

Training programs were provided by a qualified physical trainer from the Department of Sport, University College Cork. The goal was to increase physical fitness to levels necessary for participation in an endurance-based sports competition. Specifically, the two participants separately trained for a full-distance marathon and an Olympic-distance triathlon. Training for the marathon consisted of regular aerobic exercise complimented with twice weekly resistance training while preparation for the triathlon was exclusively aerobic conditioning through endurance activity. Participants met on a monthly basis with the study physical trainer to ensure graduated progression. Throughout, both participants tracked their respective physical activity with wearable activity monitors (ActiGraph wGT3X-BT, ActiGraph, FL, U.S.). These activity monitors were worn continuously on the non-dominant wrist and were removed only for cleaning, during bathing, and downloading/recharging of the monitor.

### 2.4 Anthropometric measurement visits

For the first week, participants were asked to maintain usual physical activity habits to record baseline activity levels. At each two week visit thereafter, anthropometric measurements were recorded (e.g. body mass index (BMI) and waist:hip ratio), in addition to fasting blood, fresh faecal and urine samples, and activity monitor data requisition. Participants were asked to refrain from use of alcohol, medication, and moderate to vigorous physical activity for at least 24 hours prior to each measurement visit. To minimize potential effects of diurnal variation, measurement visits took place between 7:00 a.m. and 10:00 a.m. At monthly intervals participants measured their heart rate variability using the HRV4training smartphone application [22]. Clinical variables such as blood pressure and heart rate were recorded before phlebotomy by a trained nurse. Plasma and serum samples were transported immediately to the clinical laboratories. Fresh urine and fecal samples were transported immediately at room temperature to the laboratory for DNA extraction and sample storage.

### 2.5 VO_2max_ and body composition measurement

At 0, 3, and 6 months, body composition profile and cardiorespiratory fitness (VO_2max_) were determined. VO_2max_ was determined using maximal aerobic capacity in the Astrand Treadmill test [23, 24]. This incremental protocol was conducted by an experienced exercise physiologist. The British Association of Sport and Exercise Sciences (BASES) criteria were used to define attainment of VO_2max_ [25]. VO_2max_ was achieved when i) A plateau in the oxygen uptake/exercise intensity relationship occurred (defined as an increased in oxygen uptake of less than 2 ml.kg-1.min-1); ii) A final respiratory exchange ratio of 1.15 or above was reached; iii) A final heart rate within 10 b.min-1 of the age-related maximum was reached and iv) post-exercise (5min) blood lactate of 8mmol.l-1 or more was evident.

Body composition was measured using a Lunar iDXA machine (GE Healthcare, Madison, WI) equipped with enCORE software (V.13.4, 2010) for a three-compartment body composition model (fat mass, bone mass, lean tissue). Quality control (QC) analysis was performed on the iDXA machine before use on each measurement day.

### 2.6 Inflammatory cytokine measurement

Blood samples (4 ml) from participants were collected in serum separator clot activator blood collection tubes (Greiner Bio-One, Stonehouse, United Kingdom; reference no. 454071). The blood samples were allowed to rest upright on the laboratory bench for 30 min before centrifugation at 5,000 × *g* for 10 min at room temperature. Approximately 2 ml of supernatant sera was harvested by pipette, frozen, and stored at −80°C in polypropylene cryogenic vials. Following a complete thaw, resting levels of proinflammatory cytokines were measured using a mesoscale discovery (MSD) platform (Meso Scale Discovery, Rockville, MD). The MSD system is an electrochemiluminescence-based solid-phase multiplex assay. An ultrasensitive human proinflammatory I, V-Plex immunoassay panel was used to measure serum levels of interleukin-6 (IL-6), IL-8, IL-10 and tumor necrosis factor alpha (TNF-α). Samples were diluted 1:2 according to the manufacturer’s protocol. The lower limit of detection was <1 pg/ml for all assays and standardized calibration curves were confirmed before testing. All serum samples were measured in duplicate, and the mean cytokine concentration of the duplicates (in picograms per milliliter) was used for analysis.

### 2.7 Dietary data collection

Throughout the study period participants were requested to refrain from taking vitamin, dietary, pre/probiotic, and herbal supplements, and to maintain usual *ad libitum* dietary intake. The participants were confirmed to not be taking such products at the time of recruitment. No antibiotics were used by either participant during the study period. Participants were also asked to record daily dietary habits via the MyFitnessPal smartphone application [26]. Such dietary recordings were deconstructed into macronutrient values and scaled according to average caloric intake (calories X grams/day). Dietary data were also collected by means of a monthly 146-item food frequency questionnaire (FFQ). Participants were asked to record their usual pattern of dietary intake over the previous 4 weeks. The FFQ used was an adapted version of the questionnaire used in the United Kingdom arm of the European Prospective Investigation into Cancer (EPIC) study [27], which was based on the original Willet FFQ [28].

### 2.8 DNA extraction and metagenomic sequencing of fecal samples

Biological samples (urine and fecal) were provided by participants as partial evacuations into sterile sealed containers. Upon collection samples were transported at room temperature to the Teagasc Moorepark research facility. On arrival urine samples were immediately stored at −80°C, while fecal samples were first used for DNA extraction. Sample processing and storage occurred within 6 hours of donation in the majority of cases and never after 12 hours. DNA was extracted from the donated fresh fecal samples using a QIAmp DNA stool minikit (Qiagen, Crawley, West Sussex, United Kingdom) [29]. Samples were prepared for DNA extraction by manual homogenization of the core and external surface of the fecal sample. The provided manufacturer’s protocol was enhanced using a zirconia bead (Stratech Scientific) cell disruption bead-beating step (performed three times for 30 s each time). DNA extracts and the remaining fecal samples were subsequently stored at −80°C until sequencing.

Metagenomic libraries were prepared and subsequently sequenced as previously described [2]. Briefly, libraries were generated with an Illumina Nextera XT DNA library preparation kit (Illumina Inc., USA). Normalization of library concentrations to the recommended 0.2 ng/μL was achieved with the ThermoFisher Qubit 2.0 Flurometric Quantitation system (Q32854, ThermoFisher). Following tagmentation, libraries were purified with the AMPure magnetic bead system at a ratio of 1:1.8 (DNA:AMPure) (9A63880, Beckman Coulter). An equimolar library pool of all samples was used for sequencing on an Illumina NextSeq 500 (chemistry V.2.0) sequencing platform (Teagasc sequencing facility). High-throughput sequencing was performed using the high-output 500/550 reagent kit.

### 2.9 Bioinformatic quality control of metagenomic sequencing reads

Quality control of metagenomic FASTQ sequences proceeded with the removal of host (human) reads using NCBI Best Match Tagger (BMTagger v.1.1.0). Reads were converted to Binary Alignment Map (BAM) format and sorted using FastqToSam (v.2.7.1). Low-quality reads (Phred quality score < 20), adapter sequences and short reads (Length cutoff: 105bp) were trimmed using the trimBWAstyle.usingBam.pl script. PCR duplicates were removed using MarkDuplicates from Picard tools (v.2.7.1). Finally, forward and reverse reads were merged and converted to FASTA format using IDBA fq2fa (v.1.1.1).

### 2.10 Metagenomic sequencing bioinformatic analysis

Reads which passed quality-control filtering were used as input for taxonomic profiling using MetaPhlAn2 (v.2.7.7) [30]. The top 50 most abundant species were selected for visualization. Functional profiling of high-quality processed reads was facilitated by use of the Human Microbiome Project (HMP) Unified Metabolic Analysis Network (HUMAnN2 V.0.99) pipeline [31]. MetaPhlAn2 and ChocoPhlAn pangenome database were used to facilitate fast, accurate, and organism-specific functional profiling. Models of microbial metabolic pathways were produced by HUMAnN2 which uses UniRef database to provide gene family definitions and MetaCyc provides pathway definitions by gene family. Metadata was associated with community totals using the humann2_associate package to identify altered pathways between samples. False Discovery Rate (FDR) correction for multiple tests was applied with a significance threshold of pFDR < 0.05.

### 2.11 Metabolomic sample preparation

Samples were stored at −80°C prior to analysis. Urine samples were subjected to vortex mixing and then centrifuged at 1,600 × *g* for 10 min to remove precipitated proteins and particulates. For metabolic profiling analysis by reversed-phase (RP) and hydrophilic interaction chromatography (HILIC) ultraperformance liquid chromatography-mass spectrometry (UPLC-MS), samples were prepared as follows: 200 μl of supernatant was diluted (1:1) with high-purity (ultraperformance liquid chromatography [HPLC]-grade) water, subjected to vortex mixing, centrifuged at 2,700 × *g* for 20 min, and divided into aliquots for analysis. Quality control (QC) samples were prepared by pooling 50-μl volumes of each sample. For ^1^H nuclear magnetic resonance (^1^H-NMR) spectroscopy, each sample contained 540 μl of urine mixed with 60 μl of phosphate buffer (pH 7.4; 80% D_2_O) containing a 1 mM concentration of the internal standard, 3-(trimethylsilyl)-[2,2,3,3,-2H4]-propionic acid (TSP)-2 mM sodium azide (Na^3^N), as described previously [32]. During the analyses, samples were maintained at 4°C in the autosampler.

Fecal samples underwent two freeze-thaw cycles. Following the freeze-thaw cycles, 100 mg of homogenized sample was placed in a microtube containing 250 μl of 25% acetonitrile (ACN) (1:2 ACN/H_2_O), 2 mM sodium azide, and ~0.05 g 1-mm-diameter zirconia beads. Each microtube was processed for 10 s in a Biospec bead beater. Samples were then centrifuged at 16,000 × *g* for 20 min. The fecal-water supernatant was subsequently centrifuged through centrifuge tube filters (cellulose acetate membrane; pore size, 0.22 μm) to remove any remaining particulate matter. The centrifuge tube filters were washed three times with 25% acetonitrile prior to use. The resulting fecal water was prepared for UPLC-MS profiling using HILIC by diluting 3:1 with acetonitrile and for bile acid profiling by diluting 1:1 with isopropanol. Samples were subjected to vortex mixing and incubated at −20°C for 1 h. Following the incubation step, samples were centrifuged at 4°C at 16,000 × *g* for 1 h and divided into aliquots for analysis. QC samples were prepared by pooling 20-μl volumes of each fecal-water sample followed by preparation as described above. For ^1^H-NMR spectroscopy, 50 μl of the filtered fecal water was added to a glass tube (Pyrex), which was placed under a nitrogen gas flow for 30 min or until all the liquid had evaporated. The dried sample was reconstituted with 540 μl of D_2_O and 60 μl of phosphate buffer solution as described above. The solution was mixed and sonicated for 5 min before undergoing further centrifugation at 14,000 rpm for 10 min, and then 600 μl of the supernatant was transferred to an NMR tube for ^1^H-NMR spectral acquisition.

### 2.12 Metabolomic analysis

RP, HILIC, and bile acid UPLC-MS metabolic profiling experiments were performed using a Waters Acquity Ultra Performance LC system (Waters, Milford, MA) coupled to a Xevo G2 quadrupole-time of flight (Q-TOF) mass spectrometer (Waters, Milford, MA) with an electrospray source. Samples were analyzed in randomized order, with QC analyses performed every 10 samples. First, urine samples were analyzed using UPLC-MS and an RP chromatographic method with both positive and negative MS ionization modes. Second, to separate and detect the more polar molecules, a HILIC chromatographic stage was used with the positive MS ionization mode. Fecal-water samples underwent analysis using HILIC and bile acid profiling chromatographic methods in positive and negative ionization modes, respectively. HILIC, RP, and bile acid profiling liquid chromatographic separation procedures were performed as previously described [33, 34]. Mass spectrometry was performed with the following settings. Capillary and cone voltages were set at 1.5 kV and 30 V, respectively. The desolvation gas level was set at 1,000 liters/h at a temperature of 600°C. The cone gas level was set to 50 liters/h. The source temperature was set to 120°C. To ensure the accuracy of the mass data, a lock-spray interface was used, with leucine enkephalin (556.27741 Da ([MH]), 554.2615 Da ([M–H]–)) solution used as the lock mass at a concentration of 2,000 ng/ml and a flow rate of 15 μl/min.

^1^H-NMR spectroscopy was performed on the aqueous-phase extracts at 300 K on a Bruker 600-MHz spectrometer (Bruker Biospin, Germany) using a standard one-dimensional (1D) pulse sequence corresponding to RD − *g*_z1_ − 90° − *t*_1_ − 90° − *tm* − *g*_z2_ − 90°− ACQ [32], where the value of 90° represents the applied 90° radio frequency pulse; the relaxation delay (RD) was set at 4 s, the interpulse delay (*t*_1_) was set at 4 μs, the mixing time (tm) was set at 10 ms, the magnetic field gradients (*g*_z1_ and *g*_z2_) were applied for 1 ms, and the acquisition period (AQA) was 2.7 s. Water suppression was achieved through irradiation of the water signal during RD and *tm*. Urine sample spectra were acquired using 4 dummy scans followed by 32 scans whereas fecal spectra were acquired using 256 scans and 4 dummy scans and collected into 64 K data points. A spectral width of 12,000 Hz was used for all the samples. Prior to Fourier transformation, the free induction decay (FID) values were multiplied by an exponential function corresponding to a line broadening of 0.3 Hz.

### 2.13 Metabolomic data treatment

The raw mass spectrometric data acquired were preprocessed using xcms in R. Centwave peak picking methods were used to detect chromatographic peaks [35]. The xcms-centWave parameters were data set specific. Feature grouping across samples was performed using the “nearest” method within xcms. Peak filling and MinFrac (0.5), and coefficient of variation (CV) (0.3) filters were applied to the features. Data were normalized using median fold change normalization to the median dataset [36].

^1^H-NMR spectra were automatically corrected for phase and baseline distortions and referenced to the TSP singlet at δ 0.0 using TopSpin 3.1 software. Spectra were then digitized into 20 K data points at a resolution of 0.0005 ppm using an in-house MatLab R2014a (MathWorks, Inc.) script. Subsequently, spectral regions corresponding to the internal standard (δ −0.5 to 0.5) and water (δ 4.6 to 5) peaks were removed. In addition, urea spectra (δ 5.4 to 6.3) were removed from the urinary spectra. Spectra were normalized using median fold change normalization to the median spectrum [36]. Combinations of data-driven strategies, such as SubseT optimization by reference matching (STORM) [37] and Statistical TOtal Correlation SpectroscopY (STOCSY) [38], and analytical identification strategies were used to identify metabolites of interest from ^1^H-NMR data sets. Specifically, a catalogue of 1D ^1^H-NMR and 2D NMR experiments was performed using techniques such as J-RESolved spectroscopy, ^1^H-^1^H TOtal Correlation SpectroscopY (TOCSY), ^1^H-^1^H Correlation SpectroscopY (COSY), ^1^H-^13^C Hetero-nuclear Single Quantum Coherence (HSQC), and ^1^H-^13^C Heteronuclear Multiple-Bond Correlation (HMBC) spectroscopy. Finally, for those metabolites giving ambiguous data, e.g., TMAO, the metabolites were confirmed using *in situ* spiking experiments and authentic chemical standards. Semiquantification data corresponding to the identified metabolites were calculated through peak intensity measurements of the normalized ^1^H-NMR spectra using an in-house script. GC-MS data were processed using MassHunter Quantitative Analysis (Agilent Technologies, RRID:SCR_015040) software.

### 2.14 Predicted adherence to healthy diet

Objective assessment of the participants’ adherences to WHO dietary guidelines was generated by applying a validated novel mathematical tool to ^1^H-NMR urinary profiles. Dietary patterns are predicted with the tool which implements a Monte Carlo cross-validated partial least squares-discriminant analysis (PLS-DA) model derived from urinary metabolic profiles generated from an in-patient randomized controlled clinical trial. This trial required complete adherence to diets representing various degrees of completeness of WHO healthy eating recommendations in healthy participants [39]. ^1^H-NMR urinary metabolic profiles from participants were projected into the MCCV-PLS-DA model, calculating a predicted score for each point of measurement that reflected their adherence to healthy eating.

### 2.15 Quantification and statistical analysis

Statistical assessment of dissimilarity matrices (Bray-Curtis) derived from microbial data was facilitated with the adonis2 function in the vegan R package (V.2.4-3, RRID:SCR_011950) [40]. Measurements of α-diversity and calculations of relative abundances were also performed with the vegan R package using Shannon index. Relative-abundance data were generated separately for identified species within each phylogenetic domain (e.g., *Bacteria)*.

For metabolomic analysis, the resulting ^1^H-NMR and LC-MS data sets were imported into MatLab to conduct multivariate statistical analysis. Data were centered and scaled to account for the repeated-measures design and then modelled using partial-least-squares-discriminant analysis (PLS-DA) with Monte Carlo cross-validation (MCCV) [39]. The fit and predictability of the models obtained were determined and expressed as R2 and Q2 values, respectively.

## 3 Results

### 3.1 Study overview

Two male adults aged 30 and 33 years with starting BMI of 28.6 and 31.7kg/m^2^, respectively, were recruited for participation (Table 1). During the study the participants were engaged in instructor-led improvements in physical fitness aimed towards completion of endurance events i.e., full-distance marathon and Olympic-distance triathlon respectively. Both participants demonstrated incremental improvements in measurements of body composition and cardiorespiratory fitness throughout the duration of the study (Figure 2D and Table 1). BMI, waist circumference, and resting heart rate decreased for both participants while under observation. Additionally, estimated maximal oxygen consumption (VO_2max_) increased. Total body fat (%) was also reduced.

**Table 1.** Baseline measurements of participant demographic and anthropometric characteristics. Measurements presented for initial (**T_0_**) and final (**T_14_**) readings, along with the change between the two (**Δ**). BMI, body mass index; IPAQ, International Physical Activity Questionnaire; METS, metabolic equivalents; HR, heart rate; bpm, beats per minute; PA; VO_2max_, estimated maximal oxygen consumption.

| Patient characteristics | Values |  |  |  |  |  |
| --- | --- | --- | --- | --- | --- | --- |
|  | Participant 1 (Marathoner) |  |  | Participant 2 (Triathlete) |  |  |
|  | T<sub>0</sub> | T<sub>14</sub> | Δ | T<sub>0</sub> | T<sub>14</sub> | Δ |
| Age (years) | 30 | — | — | 33 | — | — |
| Height (cm) | 181 | — | — | 182 | — | — |
| Weight (kg) | 93.8 | 89.2 | **-4.6** | 104.9 | 103.4 | **-1.5** |
| BMI (kg/m<sup>2</sup>) | 28.6 | 27.2 | **-1.4** | 31.7 | 31.2 | **-0.5** |
| Waist:Hip ratio | 0.92 | 0.92 | **0.0** | 0.95 | .91 | **-0.04** |
| Body fat (%) | 25.6 | 21.7 | **-3.9** | 34.7 | 34.5 | **-0.2** |
| Fat mass (kg) | 23.9 | 19.4 | **-4.6** | 36.3 | 35.7 | **-0.6** |
| Fat mass (trunk) (kg) | 14.8 | 11.7 | **-3.1** | 20.9 | 20.4 | **-0.5** |
| Lean tissue mass (kg) | 65.6 | 65.9 | **0.2** | 64.97 | 64.2 | **-0.7** |
| Estimated VO<sub>2max</sub> (mls/kg/min) | 41.1 | 46.6 | **5.5** | 33.6 | 38 | **4.4** |
| Max HR (bpm) | 183 | 179 | **-4** | 196 | 179 | **-17** |
| Resting HR (bpm) | 69 | 50 | **-19** | 58 | 72 | **-2** |
| Systolic BP (mmHg) | 122 | 116 | **-6** | 128 | 127 | **-1** |
| Diastolic BP (mmHg) | 77 | 75 | **-2** | 87 | 72 | **-15** |
| Weekly PA (IPAQ, METS) | 891.5 | — | — | 646.5 | — | — |
| Weekly PA (IPAQ, kCals) | 1,393.7 | — | — | 1,130.3 | — | — |

**Figure 2.**
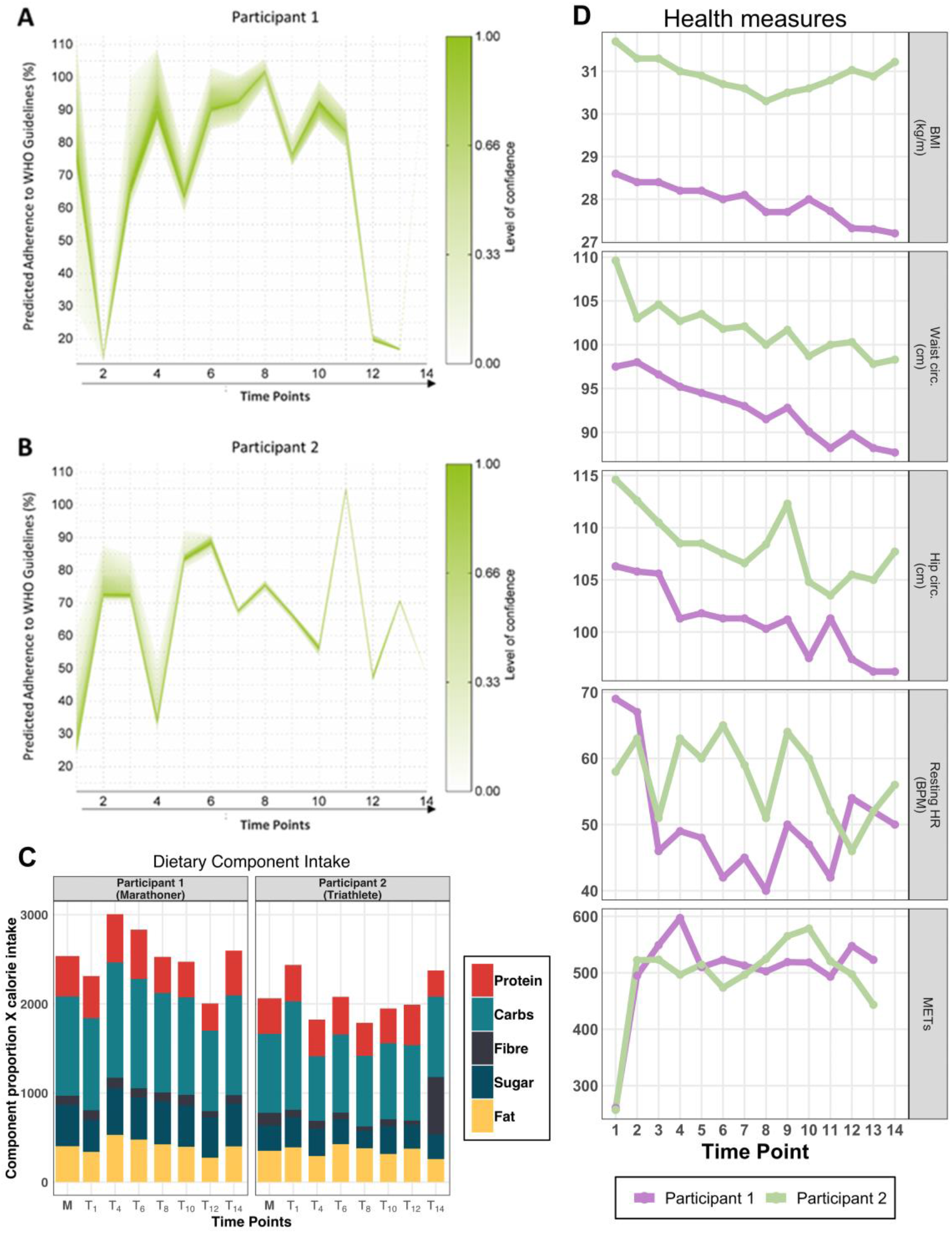
Diet and fitness characteristics. (**A** and **B**) Objective assessment of adherence to WHO dietary guidelines based on the urine composition using ^1^H-NMR urinary profiles from participant 1 (marathoner) (**A**) and participant 2 (triathlete) (**B**). The X axis represents time points of the study while the Y axis indicates the predicted percentage of adherence to WHO dietary guidelines. The level of confidence in the prediction is described by the index to the right. Participant 1 exhibits a more sustained improvement in adherence to WHO dietary recommendations (**A**). Participant 2 demonstrates more fluctuation in their adherence to WHO dietary recommendations, combined with lower levels of prediction confidence (**B**). (**C**) Nutritional constituents of participants over the study period. Daily averages of dietary macronutrients recorded over time intervals as grams per day, presented as proportions of time interval total intake, and scaled to average caloric intake per interval. Bar sums indicate average daily caloric intake, with participants’ averages for calories and dietary components indicated (M). (**D**) Participant body composition and fitness measurements. Fitness parameters improved during the observation period. BMI, hip and waist circumference, and resting heart rate (bpm) were shown to have an overall reduction. Physical activity was elevated for both participants throughout the study. BMI, body mass index; METS, metabolic equivalents; HR, heart rate; bpm, beats per minute;

Participant 1 (Marathoner) experienced the greatest improvement in cardiorespiratory fitness and body composition (Table 1 and Figure 2D). During the study the participant’s body weight was reduced by 4.6kg (4.9% of baseline), primarily corresponding to a reduction in total fat mass (−3.9%). Estimated VO_2max_ increased by 5.5mls/kg/min, while maximum and resting heart rate decreased by 4 and 19 beats per minute, respectively. Participant 2 (Triathlete) concluded the study with more modest improvements. A total body weight loss of 1.5kg (1.4% of baseline) included a loss of 0.6kg total fat and 0.7kg lean tissue mass (Table 1). Estimated VO_2max_ increased by 4.4mls/kg/min, while maximum and resting heart rate decreased by 17 and 2 beats per minute, respectively (Table 1 and Figure 2D).

Dietary recordings for both participants demonstrated that the ratios of macronutrients were consistent throughout the study, with the exception of the final recording of the triathlete, at which time considerably more fiber intake was reported (Figure 2C). To confirm the impact of diet, an objective dietary assessment was performed on metabolite composition of the participant’s ^1^H-NMR urinary profiles (Figure 2A, B). This novel mathematical model [39] generated predictions of the percentage of adherence to the WHO dietary guidelines for both participants.

Predicted healthy diet adherence demonstrated that the marathoner was engaged in healthy eating habits for the majority of the study (Figure 2A), while the triathlete showed overall improvement during the study, with all predictions at a higher percentage than baseline (Figure 2B). The study was commenced with adherence rates of approximately 80% and 30% for the marathoner and triathlete, respectively. The marathoner had a predicted percentage above 65% between time points 3 and 11, before dropping below 20% at time point 12 and 13, followed by a climb to 111% in the final time point of the study (Figure 2A). There were even more fluctuations in adherence values for the triathlete. A predicted percentage above 65% was observed between time points 5 and 9, before a spike to 104% at time point 11 and a fall to 48% at the final time point.

### 3.2 Gut microbiome response to training

Species level taxonomic profiles of the participants’ gut microbiota showed fluctuations that corresponded to reported health and training events (Figure 3). The intra-individual (α-diversity) tended to increase over the course of the study. While under observation, both participants experienced minor illnesses which were accompanied by transient reductions in diversity of bacterial species (Figure 3A). The marathoner experienced minor illness (pharyngitis, diarrhoea and wisdom tooth pain) during the 2 week period after the marathon, which was accompanied by lowered diversity of bacterial species and metabolic pathways. This reduction corresponded with an increase in IL-6 concentrations in the blood (Figure S1C). Further, this decrease in α-diversity persisted for the short remainder of the study. Similarly, a considerable drop in the triathlete’s gut microbial diversity coincided with a training reduction that resulted from a head cold, before being elevated as training resumed. Notably, bacterial and pathway α-diversity peaked for the marathoner near the date of marathon completion. The peak in the triathlete’s α-diversity coincided with a peak in his training intensity (Timepoint 10) during which he required a short course of non-steroidal anti-inflammatories for a musculoskeletal injury.

**Figure 3.**
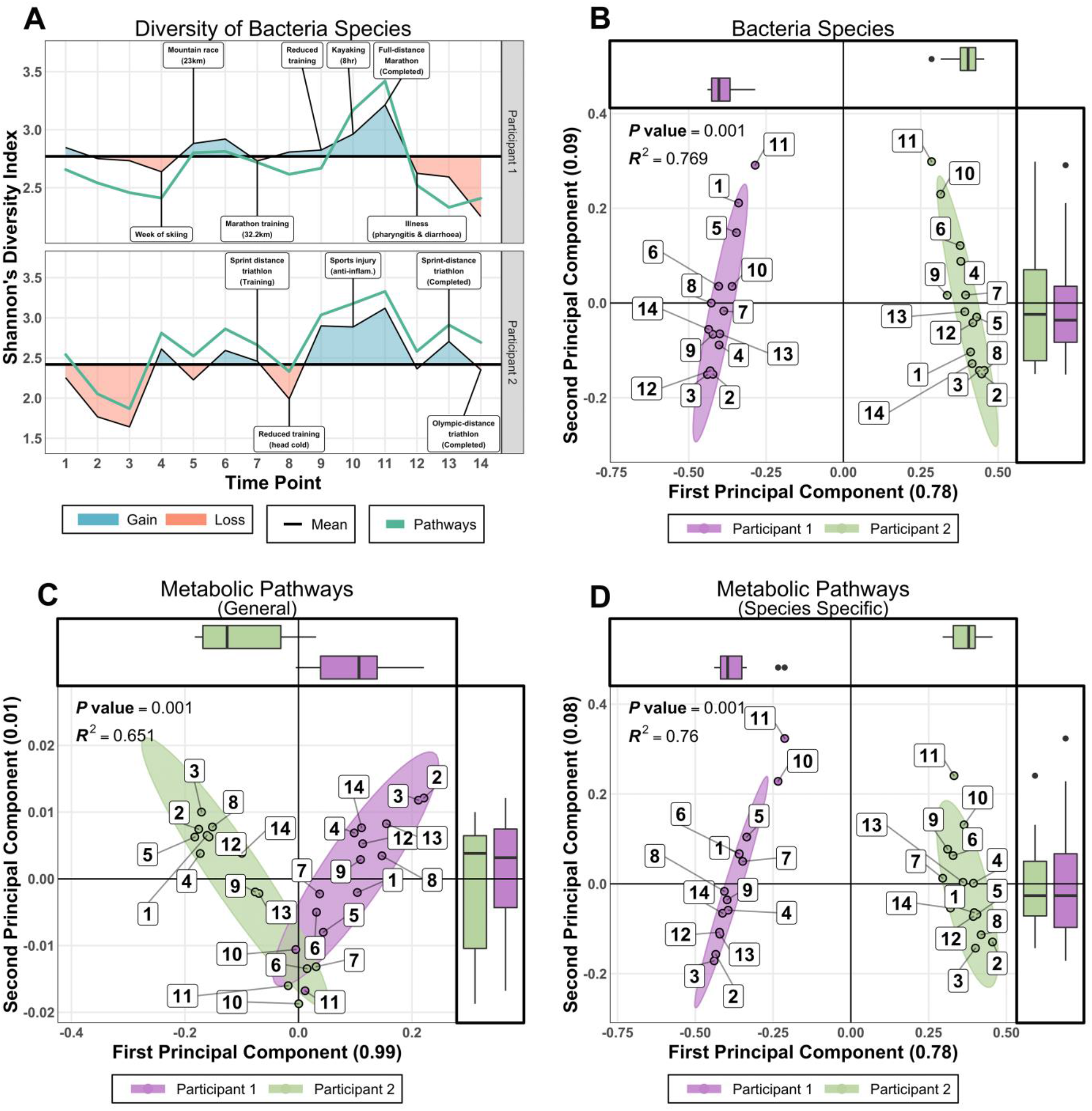
Diversity of pathways and Bacteria species. (**A**) Shannon α-diversity H-index of Bacteria species and metabolic pathway models for both participants over the duration of the study. Deviations of each participants’ mean α-diversity highlight the potential influence of training and health-related events on the composition of the gut microbiome. (**B**) β-diversity of bacterial species. PCoA of relative abundance profiles for Bacteria species demonstrates complete clustering of measurements according to individual. (**C and D**) PCoA of models for general and taxonomically related metabolic pathways. Box plots along the axes of all panels display the concentrations of data points.

Assessment of taxonomic inter-individual (β-diversity) via Principal Coordinates Analysis (PCoA) of bacteria species relative abundance illustrated considerable differences between the two participants throughout the study (Figure 3B). Differences were also observed for species specific variants of metabolic pathway models, (e.g., L-lysine degradation in *Akkermansia muciniphila*, Figure 3D). In contrast, a convergence of general pathways (e.g., L-lysine degradation, Figure 3C) was observed.

*Bacteroides* was the most abundant genus within the marathoner’s gut microbiome, with four of the top five most abundant species being *B. stercoris, B. ovatus, B. caccae*, and *B. vulgatus* (Figure 4A). For the triathlete, *Prevotella copri* was the most abundant gut bacterial species throughout the study (Figure 4B). Examination of the gradual logFC (log fold change) alteration of the most abundant species present in the marathoner highlighted a dramatic reduction in *Bifidobacterium*, *Eubacterium*, and *Roseburia* species occurring after the participant became ill. At this same point *Alistipes* species, particularly *A. senegalensis*, were elevated (Figure 4A). For the triathlete, significant reductions in the relative abundance of *Ruminococcus*, *Dorea*, and *Eubacterium* species were apparent immediately after the period of highest diversity (Figure 4B). The greatest increase observed at any time point related to *Bifidobacterium longum* (logFC = 7.6), which occurred following his greatest training increment. Further, logFC of species after 6 months training versus the initial assessment found that, for the marathoner, the relative abundance of *Veillonella parvula* increased by 3.4-fold fold, and *Agathobacter rectalis* decreased 2.6-fold (Figure S1A). For the triathlete, a 5.8-fold increase in *Methanobrevibacter smithii* and a nearly 7.4-fold decrease in *Bifidobacterium animalis* occurred, while *Akkermansia muciniphila* underwent a 2.6-fold increase in relative abundance (Figure S1B).

**Figure 4.**
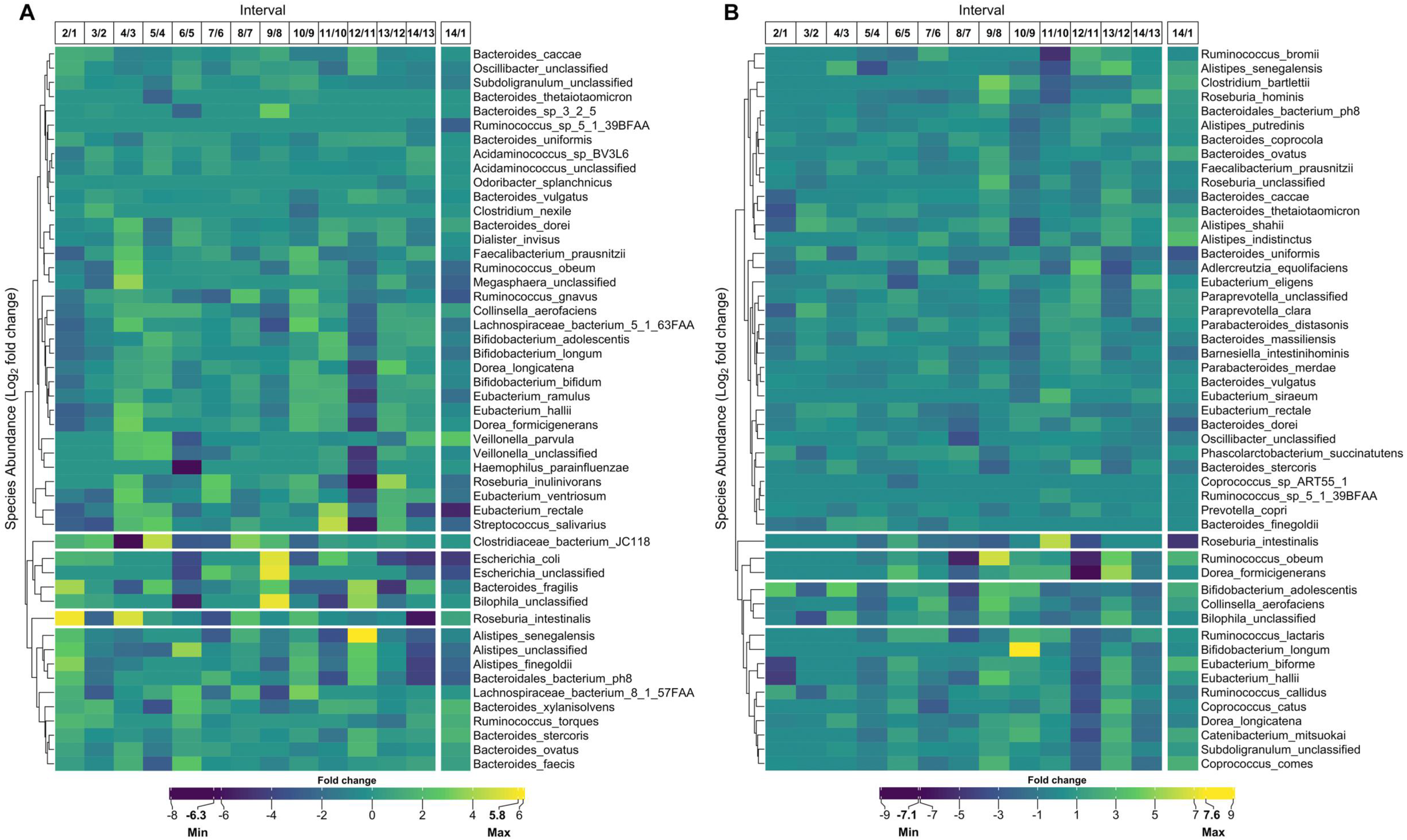
Temporal change of bacteria species. Variation in abundance of the 50 most prevalent bacteria species between sequential paired measurements and the overall change for the (**A**) marathoner and (**B**) triathlete gut microbiome. Intervals correspond to number of 2 week periods during the 6 months of training. Values are displayed as the log2 fold change of relative abundance for reported species. The clustering is performed with average linkage and Bray-Curtis distance.

### 3.3 Individual-specific metabolic changes in response to exercise over time

^1^H-NMR analysis of urine and fecal samples revealed changes in the metabolic profiles of the participants during 6 months of increased physical activity. Unsupervised PCA analysis was performed on each participant independently (Figure 5A-D). For both participants, the urinary profile scores (Figure 5A, B) demonstrated more pronounced adaptions to the exercise stimulus than was the case for fecal profiles (Figure 5C, D), as outlined by a smoother trajectory with disparate beginning and end points (0 and 26weeks).

The score plot pertaining to the urinary profile of the marathoner (Figure 5A) showed a clearer pattern than was the case for the triathlete (Figure 5B). The data points mostly moved from the upper quadrants to the lower over time, with greater correlation among points, as indicated by lower dispersal about the trajectory. The opposite was observed for the fecal profile score plots. While the marathoner’s fecal profile (Figure 5C) did not undergo distinct changes over time, the triathlete’s fecal profile (Figure 5D) underwent a distinctive shift of data points from the upper quadrants to the lower over time.

**Figure 5.**
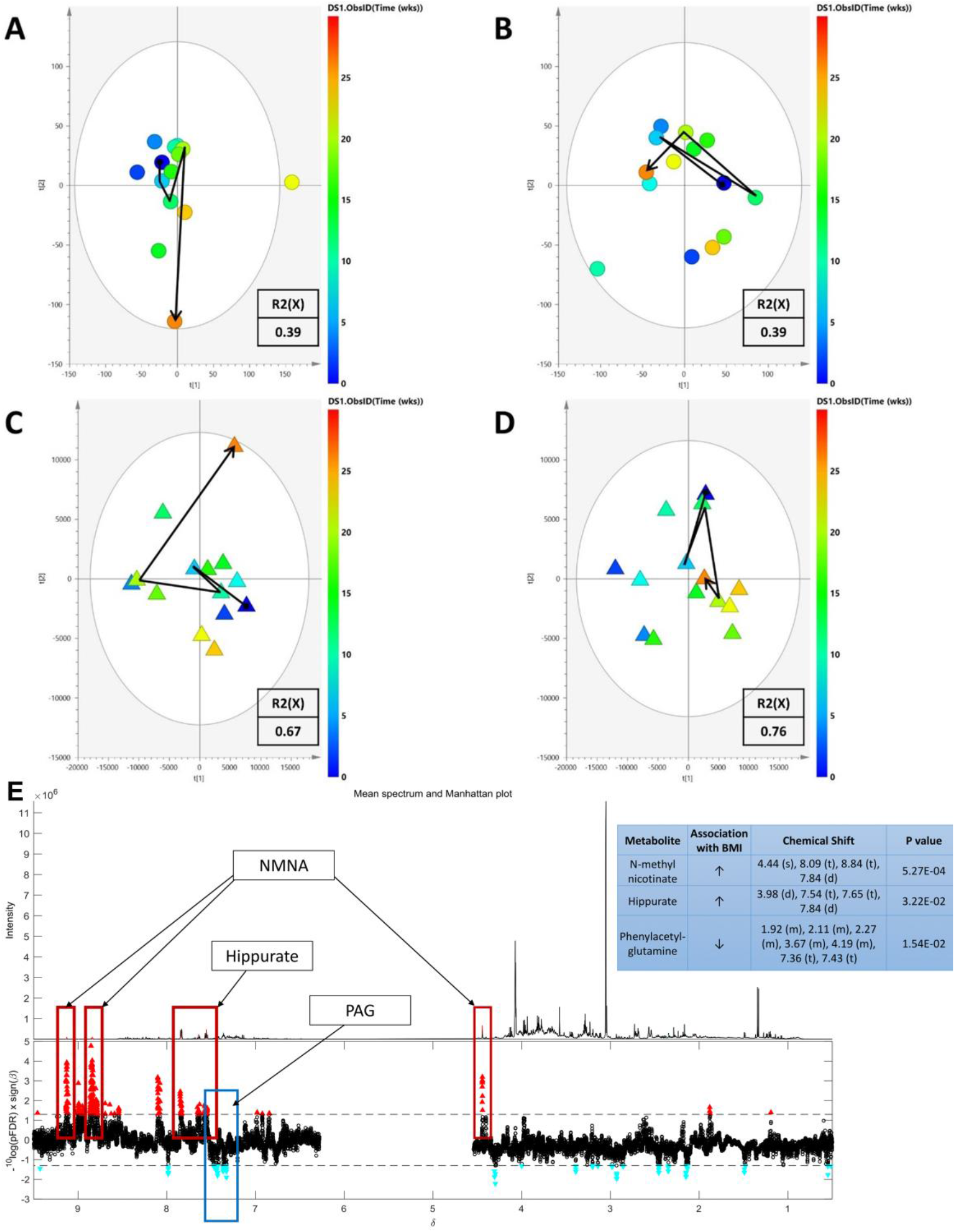
PCA analysis of ^1^H-NMR urinary and faecal datasets. PCA-time trajectory scores plot of ^1^H-NMR urine and fecal samples of the (A and C, respectively) marathoner and (B and D, respectively) triathlete. Each sample is represented by a dot which is colored according to time (weeks) where dark blue is 0 weeks and dark orange is 26 weeks. A time trajectory was constructed using data points at 0, 6, 12, 20 and 26 weeks. Urinary profiles (A and B) exhibit a more concise path over time compared to cognate fecal profiles (C and D), with the marathoner’s urinary profile demonstrating differences in the metabolic profile that are more conserved between data points (A). Ability of the model to account for variation within the dataset is described by the R2(X) values.

### 3.4 Disparate metabolic alterations in response to exercise are associated with weight loss

PCA-time trajectory plots of the ^1^H-NMR urine samples from both participants (Figure 5A, B) were labelled for BMI (Figure S2A, B), describing an association to body weight changes. PCA time trajectories were further color-coded according to various read-outs of fitness and diet in order to compare progress of the participants over the six months of increased physical activity (Figures S3, S4). The participants demonstrated similar patterns for cardiorespiratory fitness, (VO_2max_, Figures S3C, S4C) and reported consumption of fruits and vegetables (Figures S3F, S4F), while differences were observed for step count (Figures S3A, S4A), moderate exercise duration (Figures S3B, S4B), protein consumption (Figures S3D, S4D), and fiber intake (Figures S3E, S4E).

### 3.5 Exercise responsive metabolites linked with BMI

To explore the relationship between exercise and body habitus, we identified the metabolites significantly associated with BMI within the profiles of both participants. Given the greater reflection of metabolic changes within the urinary datasets, linear regression was performed on the combined urinary profiles of the two participants, with BMI as the independent variable. Three metabolites were significantly associated to BMI (pFDR < 0.05). N-methyl nicotinate (NMNA) and hippurate were shown to be positively correlated with BMI, while phenylacetylglutamine (PAG) was shown to be inversely related with BMI (Table 2). The UPLC-MS combined urinary datasets of both participants were also subject to linear regression to BMI in order to discover any associated amino acids. L-Serine, L-Asparagine, L-Isoleucine and D-2-aminobutyric acid were identified as being negatively correlated with BMI (Table 2).

**Table 2.**
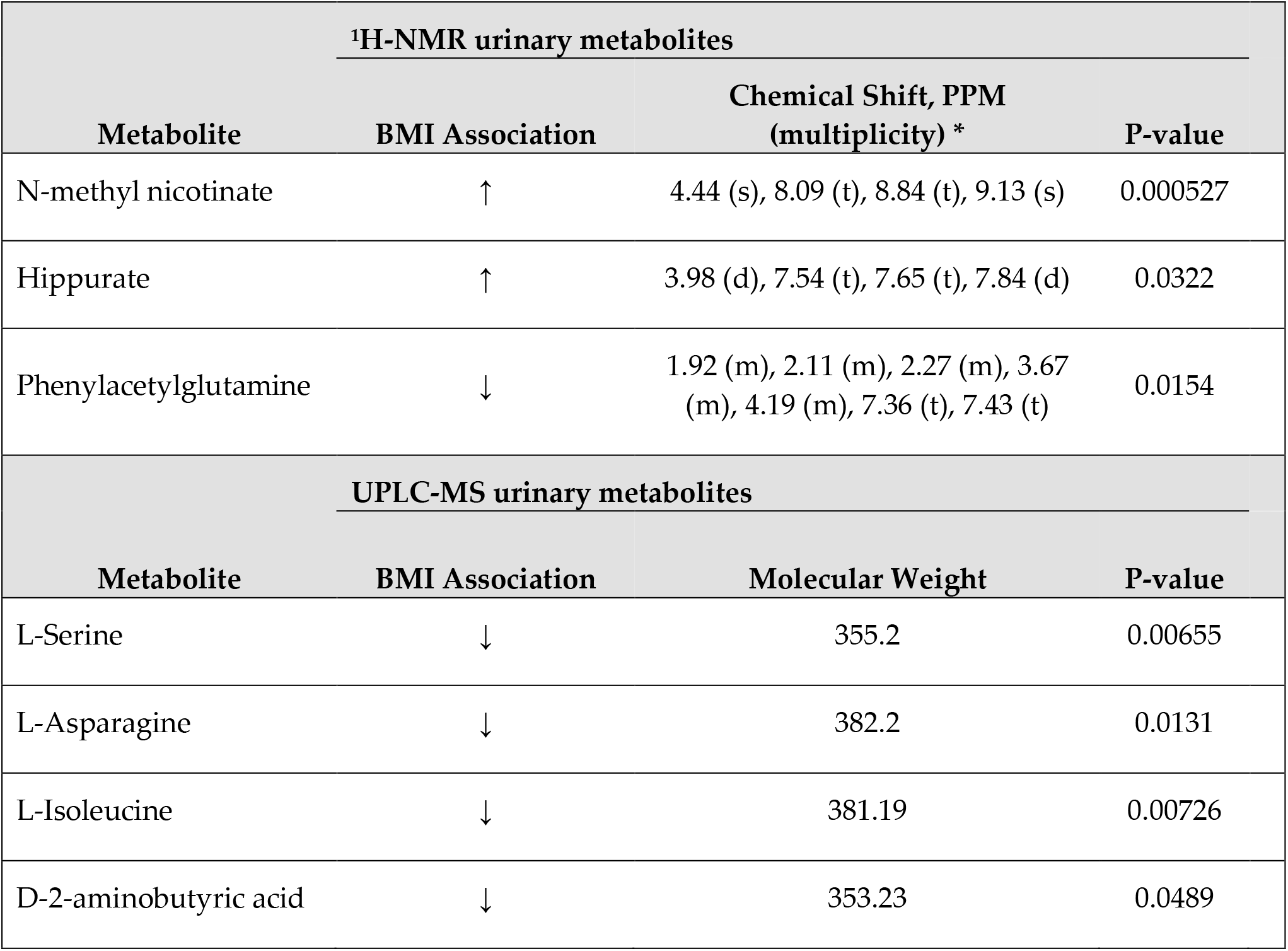
Significant metabolites identified through linear regression to BMI for both volunteer’s ^1^H-NMR and UPLC-MS urinary datasets. N-methyl nicotinate (NMNA) and hippurate were shown to be positively correlated with BMI while phenylacetylglutamine (PAG) demonstrated inverse correlation with BMI. All significant UPLC-MS metabolites were negatively correlated with BMI. For ^1^H-NMR profiles, chemical shifts of each metabolite, as well as their corresponding multiplicity were used in the identification process. A significant p-value threshold of 0.05 was chosen after calculating the false discovery rate (FDR)

## 4 Discussion

Our prior investigations characterizing the gut microbiome within the framework of physical exercise have included profiling extremes of fitness, as assessed through examination of the elite athlete gut microbiome [12, 41], and also investigating the impact of structured exercise in the physically inactive [1, 2]. Although profiling of the athlete gut microbiome revealed numerous differences in taxonomic composition, functional potential, and metabolic activity relative to a control population of healthy but less fit adults, the microbiome alterations resulting from a short-term exercise intervention were more nuanced. We have theorized that the structure of the athlete microbiome is, in part, the result of adaptations to long-term engagement in rigorous physical activity and associated lifestyle (e.g., optimized diet) [42]. This is consistent with the observation that the gut microbiome of adults has been shown to be resistant to dramatic alteration and, thus, may not rapidly adapt to the systemic influences of exercise [43].

Here however, we investigated whether the gut microbiome is altered by exercise over an extended period of time. Case studies are well suited to such questions but have been under-utilized in microbiome science. By tracking two participants over a 6-month period of fitness improvement, modification of the gut microbiome in response to well-documented training events, dietary trends, and health events was achieved.

The diversity of gut bacterial species increased with physical training, while also responding to periods of injury and illness. Measurements of α-diversity for the marathoner peaked at the sample collection point nearest to the date of the marathon and took approximately 20 weeks of sequential training to manifest a sustained increase. Likewise, the level of diversity for the triathlete peaked with a period of increased training activity, which subsequently resulted in injury and short-term non-steroidal anti-inflammatory use. It has previously been shown that a wide range of pharmaceutical compounds can interact with components of the intestinal microbiome [44, 45], but we believe that this short term medication use was unlikely to account for the concurrent increase in alpha diversity. Changes observed in the abundance of bacterial species from the participants illustrated that the microbial communities of the two individuals were distinct from one another. Despite this, between the two participants, taxa such as *Lachnospira, Dorea, paraprevotella, Faecalibacterium*, and *Ruminococcus*, experienced changes in relative abundance that have been identified in other similar studies [4, 14]. However there are disparities between these studies that are likely due to diet, study population, and exercise type. The dominance of *Bacteroides* in the profile of the marathoner did not mirror characterizations of gut microbiota in elite athletes, in which high abundance of *Bacteroides* is uncommon [13, 41]. However, an examination of soldiers undergoing intense physical military training demonstrated a decrease of *Bacteroides* that was inversely related to intestinal permeability [46]. Additionally, a recent study found that a high *Prevotella-Bacteroides* ratio reflected decreased body fat [47], which is consistent with the findings presented here. It is also interesting to note the lower degree of fold-changes across species in the marathoner compared to the triathlete. This relative stability suggests that *Bacteroides* dominant gut microbiomes may be more resilient to exercise-related perturbations. Further, the relative abundance of *Veillonella parvula* underwent a 3.4-fold increase. *V. parvula* is part of the normal microbiota of the mouth, and has been implicated in oral infections [48]. Here, the marathoner contracted pharyngitis and had wisdom tooth pain in the 2 weeks after the marathon. However, it is also notable that a recent examination of the gut microbiota of marathon runners discovered an increase in *Veillonella* species abundance post marathon [15]. The authors of that study conducted transplantation of *V. atypica* to mice, resulting in an enhancement of running endurance, which was theorized to result from improved lactate metabolism by the species. Specifically, *V. atypica* converts lactate to propionate, a short-chain fatty acid (SCFA) with numerous biological functions, including acting as a significant energy source. It may be the case that other *Veillonella* species facilitate similar metabolic function, and increased abundance of such species is a natural adaptive response to environmental changes in the gut resulting from increased physical activity.

A high abundance of *Prevotella copri* was noted within the triathlete’s gut microbiota. *Prevotella* has previously been shown to be abundant in elite cyclists performing endurance training [13], although Prevotella-dominated microbiota have also been associated with rheumatoid arthritis disease progression [49]. Robust examination of > 1000 *P. copri* genomes has described multiple distinct clades for the taxon with substantial functional variation [50]. Conceivably, this improved characterization of *P. copri* will enable more accurate determination of its role in physical fitness and other aspects of health. *Methanobrevibacter smithii* was the species that most considerably increased in relative abundance in response to training in the triathlete (5.8-fold). *M. smithii* has previously been identified in a study of cyclists at different competitive levels. The authors described that *M. smithii* was found primarily in the most competitive participants, and that high methane metabolism associated with the microbe had broad implications with other metabolic processes, including those related to SCFAs. Additionally, *M. smithii* has been shown to be increased in Anorexia Nervosa patients [51, 52] and in rats with eating restrictions [53]. The triathlete averaged ~2,000 calories per day throughout this training period, whereas the marathoner averaged ~2,500 calories per day. The combination of low-calorie diet and increased aerobic physical activity may be associated with the increase in *M. smithii* abundance.

Assessment of β-diversity for both bacterial species and taxonomically linked metabolic pathway profiles demonstrated that the participants maintained distinct microbial structures throughout the observation period. Conversely, the participants’ general metabolic pathway profiles converged for a period of the study. This suggests that broad alterations of the microbiome in response to exercise do not result in a specific structure of the microbial community, but rather that targeted adaptations occur with microbial metabolic activity. Despite this, metabolomic analysis also identified trajectories of metabolite profiles for the two participants that were dissimilar, possibly reflecting the differences exercise training pursued by the two volunteers. Curiously, significant associations between urinary metabolites and BMI were detected. One such metabolite, PAG, has previously been associated with lean body composition [54]. This association is also highlighted here, as levels of urinary PAG decrease with BMI.

It is important to note that while still poorly understood, the mechanisms underpinning cross talk between exercise and gut microbiota are beginning to be described [9]. The depletion of gut microbiota in mice resulted in decreased running endurance, which was recovered following replacement of the microbiota [55]. Similar work demonstrated significant reductions in SCFAs, gut microbiota, and endurance capacity with antibiotic treatment [56]. Infusion of the SCFA acetate resulted in the restoration of endurance capacity.

Previous investigations have established enrichment of SCFAs in elite athletes [12], however this has not been widely observed in other populations engaged in exercise, including the participants presented here. Despite dramatic changes in SCFA profile not being detected, the participants exhibited increased abundance of microbial species that have been shown to influence SCFA production. It is conceivable that enrichment of these and other related microbes precedes pronounced metabolomic alteration, and that such changes were in the process of developing. Alternatively, it may be that these microbes require substrates that were not sufficiently provided by the participants’ respective diets.

## 5 Conclusion

These results provide further evidence that the human gut microbiome is affected by exercise. We observed an increase in alpha diversity after 3-4 months of sustained and incremental training. This did not appear to differentiate between training regimens and is encouraging that regular training, if sustained, can lead to favourable microbiome characteristics. We observed a stable macronutrient intake in both case studies, suggesting that dietary change is not accountable for the observed changes in this study. However, the effects fluctuated in response to environmental stressors, such as reduced training following injury and in particular, illness. Alterations in taxa and metabolite profiles occurred in response to training, and in many instances mirror changes seen in other exploratory studies. Despite this, the current investigation it is not without limitation. While prospective dietary recording by participants via the chosen method is arguably more sensitive than retrospective recall, it still remains prone to error, if for instance food items are omitted or substance quantity miscalculated. Consideration of dietary influence should remain mandatory in the reporting of studies in this field and investigators should continue to strive for more accurate information.

The low sample size of the current study is also acknowledged, preventing generalisability to the general population, and also limits advanced statistical analysis of the gut microbiome and metabolome. The choice of a case study design was purposeful however, facilitating a real-world, longitudinal permitting interpretation and observation of the outcomes of interest in the context of individual traits and environmental incidents.

## Data Availability

The microbial DNA sequences generated for this study have been deposited in the European Nucleotide Database (ENA) database under ID code PRJEB27624.

## 7 Author Contributions

FS, MGM, and OC conceived of the study design. OC and TW formulated and supervised the physical training programs. OC managed study recruitment, enrollment, and clinical visits. CM conducted and reported on the DEXA scans for this study. WB conducted fecal DNA extraction and metagenomic library preparation. OO, RW, and WB processed sequencing data and performed subsequent microbiome analysis. IG-P and EH performed sample processing and analysis of metabolomics data. OC, WB, RW, OO, IG-P, PDC, and FS wrote the manuscript. All authors had access to the study data and critically reviewed, revised, and approved the final manuscript.

## 9 Acknowledgments

The authors would like to acknowledge Ms. Helena Nugent who assisted with volunteer measurement throughout the study and Yvonne McCarthy Department of Medicine UCC, for performing cytokine measurement. We thank the participants for their involvement in the trial.

